# Polygenic Risk of Epilepsy and Post-Stroke Epilepsy

**DOI:** 10.1101/2023.09.18.23295739

**Authors:** Cyprien A. Rivier, Santiago Clocchiatti-Tuozzo, Shubham Misra, Johan Zelano, Rajarshi Mazumder, Lauren H Sansing, Adam de Havenon, Lawrence J. Hirsch, David S. Liebeskind, Emily J. Gilmore, Kevin N. Sheth, Jennifer A. Kim, Bradford B. Worrall, Guido Falcone, Nishant K. Mishra

## Abstract

**Background and Aims:** Epilepsy is highly heritable, with numerous known genetic risk loci. However, the genetic predisposition’s role in post-acute brain injury epilepsy remains understudied. This study assesses whether a higher genetic predisposition to epilepsy raises post-stroke or Transient Ischemic Attack (TIA) survivor’s risk of Post-Stroke Epilepsy (PSE).

**Methods:** We conducted a three-stage genetic analysis. First, we identified independent epilepsy-associated (*p*<5x10^−8^) genetic variants from public data. Second, we estimated PSE-specific variant weights in stroke/TIA survivors from the UK Biobank. Third, we tested for an association between a polygenic risk score (PRS) and PSE risk in stroke/TIA survivors from the All of Us Research Program. Primary analysis included all ancestries, while a secondary analysis was restricted to European ancestry only. A sensitivity analysis excluded TIA survivors. Association testing was conducted via multivariable logistic regression, adjusting for age, sex, and genetic ancestry.

**Results:** Among 19,708 UK Biobank participants with stroke/TIA, 805 (4.1%) developed PSE. Likewise, among 12,251 All of Us participants with stroke/TIA, 394 (3.2%) developed PSE. After establishing PSE-specific weights for 39 epilepsy-linked genetic variants in the UK Biobank, the resultant PRS was associated with elevated odds of PSE development in All of Us (OR:1.16[1.02-1.32]). A similar result was obtained when restricting to participants of European ancestry (OR:1.23[1.02-1.49]) and when excluding participants with a TIA history (OR:1.18[1.02-1.38]).

**Conclusions:** Our findings suggest that akin to other forms of epilepsy, genetic predisposition plays an essential role in PSE. Because the PSE data were sparse, our results should be interpreted cautiously.

## INTRODUCTION

Post-stroke epilepsy (PSE) is a debilitating complication of stroke associated with poor cognition, functional outcomes, and increased mortality.^1^ Its incidence ranges from 2-20% and increases to 50% in the elderly population over 60 years.^2^ The epilepsy risk is 23 times greater after a stroke than in the general population.^3^ Although several risk factors for PSE have been identified,^4^ the underlying mechanisms of this condition remain poorly understood.

Genetic factors account for approximately 30% of epilepsy syndromes.^5^ Genome-wide association studies (GWAS) have identified common genetic risk variants associated with various forms of epilepsy.^6,7^ Genetic polymorphisms have been suggested to play a role in PSE development.^8^ However, the current evidence is derived from small observational studies investigating individual genetic variants with a small effect size that cannot predict risk or inform prognosis or treatment individually.^9-12^ A spectrum of genetic factors may contribute to PSE development, including rare mutations with significant effects, uncommon variants with moderate effects, and common variants with minor risk effects.

Polygenic risk scores (PRS) are a promising tool designed to measure the combined effects of several variants into a single score, each with a small impact on susceptibility, and stratify individuals based on their risk of developing the condition.^13^ Recent studies suggest that patients with high genetic predisposition to a given disease, as measured through PRS, have a 3- to 5-fold increased risk of developing the condition, similar to the risk conferred by rare monogenic variants in other diseases.^14^ PRS have been utilized to predict the risk of developing various neurological conditions,^15,16^ including epilepsy,^17^ and may have the potential for predicting the risk of PSE.

Therefore, this study aims to investigate the association between polygenic predisposition to epilepsy and PSE risk in stroke/transient ischemic attack (TIA) survivors.

## METHODS

### Study Design

Our research involved a three-stage genetic association analysis. The first stage entailed the selection of independent genetic risk variants related to epilepsy using publicly available resources. The second stage comprised estimating PSE-specific variant weights within the UK Biobank (UKB). In the third stage, we evaluated the PSE-specific PRS within All of Us. For the second and third stages, our primary analysis included all ancestries, while a secondary analysis was restricted to European ancestry only. Additionally, we performed a sensitivity analysis including only participants with stroke.

Our analytical samples for the UKB and All of Us comprised participants with both genetic data available and validated diagnoses of stroke or TIA during both study durations.

### Data Source

We used individual-level data from both the UKB and All of Us studies. The UKB is a large population-based cohort study that enrolled 502,480 community-dwelling persons aged 40 to 69 years in the United Kingdom at recruitment between March 2006 and October 2010.^18^ Study participants underwent multiple baseline physical measures, provided blood, urine, and saliva samples for different analyses, provided detailed information about themselves, and consented to monitor their health.^18^ Similarly, the National Institutes of Health’s All of Us Research Program^19^ is an ongoing study in the US that aims to enroll 1 million Americans aged 18 and older. All of Us does not focus on any particular disease or health status and seeks to recruit groups that have been underrepresented in biomedical research.^19^

Their respective institutional review boards (IRB) approved the protocols for the UKB study and All of Us (https://www.ukbiobank.ac.uk/learn-more-about-uk-biobank/about-us/ethics and https://allofus.nih.gov/about/who-we-are/institutional-review-board-irb-of-all-of-us-research-program). All participants or their legally designated surrogates provided written informed consent.

### Ascertainment of clinical phenotypes

Within the UKB, stroke diagnoses were obtained through a validated diagnostic algorithm combining baseline health surveys, electronic health records, and death registries.^20^ Diagnoses of TIA and epilepsy were made using the corresponding International Statistical Classification of Diseases and Related Health Problems, Tenth Revision (ICD-10) codes: G45 and G40.^21-23^ The information regarding stroke subtypes was only available for some stroke diagnoses; thus, all stroke subtypes were considered in the UKB. The All of Us dataset used equivalent ICD-9, ICD-10, and Systematized Nomenclature of Medicine (SNOMED) codes to ascertain ischemic stroke, TIA, and epilepsy (Supplemental Table 1).

### Genomic Data in the UK Biobank and All of Us Studies

In the UKB, genotyping was carried out by the Affymetrix Research Services Laboratory using the UKB Axiom Array. Principal component analysis was used to evaluate and assign ancestry. This pipeline yielded 488,377 samples and 805,426 markers that entered the imputation process, which was completed using the IMPUTE4 software (University of Oxford) and a combination of 3 reference panels: Haplotype Reference Consortium, UK10K haplotype, and 1000 Genomes Phase 3.^24^ The imputation process resulted in a data set with 93,095,623 autosomal single nucleotide polymorphisms (SNPs), short indels, and large structural variants in 487,320 participants. All of Us is collecting biospecimens and generating genomic data for all participants who have consented among its participants. Genome centers established a consistent sample and data processing protocol for array and WGS data generation to attenuate the likelihood of batch effects across centers. The protocol for library construction was the PCR Free Kapa HyperPrep, the sequencer used was the NovaSeq 6000, and the software used was DRAGEN v3.4.12. The software produces the metrics that are consumed by the sample QC processes. Genome centers generate variant calls submitted to the Data and Research Center. The genome centers use the same lab protocols, scanners, software, and input files.^25^

### Genetic risk variants for epilepsy

In the first analysis stage, we leveraged the GWAS Catalog to ascertain the most significant variants associated with epilepsy. This analysis yielded 39 common, independent (R2<0.01) SNPs with a minor allele frequency exceeding 5% and demonstrating a significant association with focal or generalized epilepsies at the genome-wide level (*p*<5x10^−8^) (Supplemental Table 2). Then, we adjusted the effect estimates of these SNPs based on their association with the risk of PSE in stroke/TIA survivors, as estimated in the UK Biobank. These effect estimates were derived from multivariable logistic regression models, controlling for age, sex, and the first four genetic principal components. In the third analysis stage, undertaken within the All of Us dataset, we identified proxy variants in high linkage disequilibrium (LD >0.9) for 20 of the 39 SNPs, which were not present in the All of Us genomic information. A proxy SNP, due to its close genomic proximity and thus high coinheritance with the SNP of interest, can serve as an effective stand-in when the latter is unavailable.

### Statistical Analysis

In the second stage, we applied multivariable logistic regression models, incorporating age, sex, genetic ancestry, and the first four genetic principal components to determine the association between the 39 epilepsy-related SNPs and PSE among stroke/TIA survivors in the UK Biobank. In the third stage, we used the genetic risk variants identified in Stage 1 and the PSE-specific weights determined in Stage 2 to develop a PRS that models the polygenic risk to PSE. We used univariable and multivariable logistic regression analyses to test for association between the PSE-specific PRS and PSE among stroke/TIA survivors within All of Us. The PSE-specific PRS was modelled as a continuous variable and standardized by subtracting by its mean and dividing by its standard deviation.

#### Sensitivity analysis

The analysis was replicated in stroke survivors only, excluding participants who suffered from a TIA. For this analysis, the regression models used in the second and third stages were adjusted for age, sex, genetic ancestry and the first four genetic principal components.

#### Secondary analyses

The analysis was replicated in participants of European ancestry. For this analysis, the regression models used in the second and third stages were adjusted for age, sex, and the first four genetic principal components. In addition, to verify that we constructed a PSE-specific PRS instead of a non-specific epilepsy PRS, we evaluated whether the PSE-adjusted PRS was more predictive of PSE among stroke/TIA survivors than epilepsy in the general population. Accordingly, we executed multivariable logistic regression analyses in All of Us, treating epilepsy as the outcome variable, the PSE-specific PRS as the exposure variable, and age, sex, genetic ancestry, and the first four genetic principal components as covariates.

We used the R statistical package version 4.2 for all statistical analyses and PLINK 1.9 for genetic analyses.

## RESULTS

In the UK Biobank, we identified 19,708 participants (mean age: 61.1, 56.9% female) with a stroke/TIA history. Among them, 805 (4.1%) were subsequently diagnosed with epilepsy. In All of Us, out of 12,251 participants (mean age: 64.5, 54.5% female) who suffered from a stroke/TIA, 394 (3.2%) were later diagnosed with epilepsy. The baseline characteristics of both cohorts are detailed in Table 1. In the first stage, we identified 39 independent epilepsy-linked genetic risk variants at genome-wide levels (*p*<5x10^−8^) (see Supplemental Table 2). In stage 2, we estimated PSE-specific weights for independent genetic risk variants in Supplemental Table 2. In stage 3, we focused on PSE-specific PRS and PSE in All of Us, the univariable analysis showed that a 1 SD increase in the PSE-specific PRS was associated with a 13% increase in the odds of developing PSE (OR 1.13; 95% CI 1.02 – 1.24; *P*-value= 0.01). After accounting for age, sex, race and ethnicity, and principal components, we found a 16% increase in the risk of developing PSE for a 1 SD increase in the PSE-specific PRS (OR: 1.16, 95%CI: 1.02-1.32; *P*-value 0.01 – Table 2).

**Table 1:** Characteristics of the two cohorts.

|  | UK Biobank | All of Us |
| --- | --- | --- |
| **Baseline demographics** |  |  |
| Number of participants (n) | 19,708 | 12,251 |
| Age (mean (SD)) | 61.08 (6.65) | 64.48 (13.01) |
| Female sex | 11,205 (56.9%) | 6,661 (54.4%) |
| European ancestry | 16,903 | 5,953 (48.6%) |
| **Events of interest** |  |  |
| Stroke | 12,665 (64.3%) | 8,413 (68.7%) |
| TIA | 9,415 (47.8) | 6,076 (49.6%) |
| Epilepsy | 1,334 (6.8%) | 760 (6.2%) |
| Post-stroke epilepsy | 805 (4.1%) | 394 (3.2%) |

**Abbreviations-** SD: Standard Deviation; TIA: Transient Ischemic Attack; NA: Not Available.

**Table 2:**
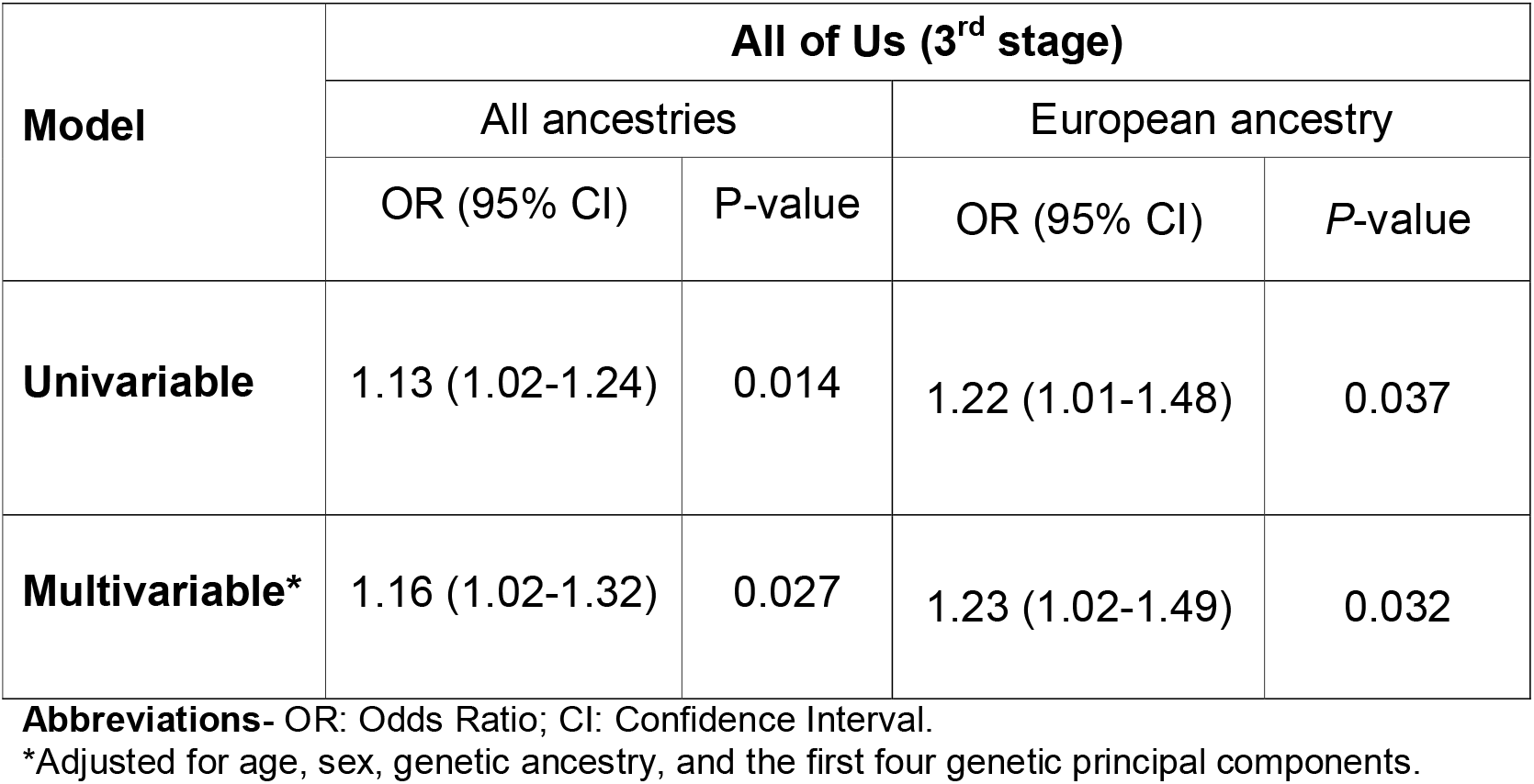
Odds Ratios of developing PSE for one standard deviation increase in the PSE-specific PRS.

| Model | All of Us (3<sup>rd</sup> stage) |  |  |  |
| --- | --- | --- | --- | --- |
|  | All ancestries |  | European ancestry |  |
|  | OR (95% CI) | P-value | OR (95% CI) | P-value |
| **Univariable** | 1.13 (1.02-1.24) | 0.014 | 1.22 (1.01-1.48) | 0.037 |
| **Multivariable\*** | 1.16 (1.02-1.32) | 0.027 | 1.23 (1.02-1.49) | 0.032 |

**Abbreviations-** OR: Odds Ratio; CI: Confidence Interval.
\*Adjusted for age, sex, genetic ancestry, and the first four genetic principal components.

### Sensitivity Analysis

When the analysis was restricted solely to participants with a history of stroke and excluded participants with TIA, the findings were consistent with our primary results. Out of the 3,714 participants who had experienced a stroke, 104 (2.8%) later developed epilepsy. Upon adjusting for age, sex, and principal components, an 18% increase in the risk of developing PSE was observed for every 1 SD increment in the PSE-specific PRS (OR: 1.18, 95% CI: 1.02-1.38; *P*-value: 0.03 – Supplementary Table 3).

**Table 3:** Odds Ratios of developing epilepsy for one standard deviation increase in the PSE-specific PRS.

| Model | All of Us (3<sup>rd</sup> stage) |  |  |  |
| --- | --- | --- | --- | --- |
|  | All ancestries |  | European ancestry |  |
|  | OR (95% CI) | P-value | OR (95% CI) | P-value |
| **Univariable** | 1.05 (1.02-1.09) | 0.002 | 1.05 (1.02-1.09) | 0.002 |
| **Multivariable\*** | 0.99 (0.95-1.03) | 0.571 | 0.99 (0.95-1.03) | 0.639 |

**Abbreviations-** OR: Odds Ratio; CI: Confidence Interval.
\*Adjusted for age, sex, genetic ancestry, and the first four genetic principal components

### Secondary Analyses

The analysis restricted to participants of European ancestry yielded similar results. Of 5,953 participants with a stroke/TIA history, 155 (2.6%) subsequently developed epilepsy (Supplemental Table 4). After accounting for age, sex, and principal components, we found a 23% increase in the risk of developing PSE for a 1 SD increase in the PSE-specific PRS (OR: 1.23, 95%CI: 1.02-1.49; *P*-value 0.03 – Table 2).

We also verified that the constructed PRS was PSE-specific. Among 312,924 All of Us participants with genetic data, 3,684 (1.2%) had epilepsy. Multivariable logistic regression results comparing the PSE-specific PRS against epilepsy were non-significant (OR: 0.99, 95%CI: 0.95-1.03; *P*-value=0.57 – Table 3).

## DISCUSSION

The present study aimed to investigate the association between polygenic predisposition to epilepsy and PSE risk in stroke/TIA survivors using a PRS approach. Our results demonstrate that a PRS constructed from epilepsy GWAS loci was significantly associated with PSE risk in both the UK Biobank and All of Us cohorts, indicating that genetic factors play an important role in developing PSE in stroke/TIA survivors. We also identified that the PSE-specific PRS was significantly associated with epilepsy risk in stroke survivors excluding TIA, and not significantly associated with epilepsy risk in the general population, indicating that it is specifically related to the risk of developing epilepsy in individuals with a stroke/TIA history.

The results of this study advance our understanding of the genetic mechanisms underlying PSE^26^ and lays the groundwork for designing future studies to characterize individuals at high PSE risk. An advanced understanding of the genetic contributions to PSE risk may inform our understanding of the therapeutic targets for its prevention and treatment. Our study findings are consistent with previous studies showing that genetic factors’ contribute to the PSE development ^8,27^ and that a PRS can predict the risk of developing epilepsy.^17,28^

Our study is the first to construct a PSE-specific PRS using GWAS-identified variants and demonstrate its association with PSE risk in two large stroke/TIA cohorts. The use of PRS provides a way to capture the collective genetic effects of multiple variants and quantify an individual’s genetic risk for a specific disease or trait.^13^ In All of Us, a 1 SD increase in PSE-specific PRS was associated with 16% higher PSE odds; similarly European ancestry saw a 23% increase. This implies the polygenic epilepsy risk may contribute to PSE in stroke/TIA survivors, providing new insights into the underlying mechanisms of this condition.

Moreover, our study highlights the potential utility of PRS for predicting PSE risk in stroke/TIA survivors. Our sensitivity analysis, which included only participants with a history of stroke and excluded those with TIA, yielded consistent results. This further supports the robustness of our findings and emphasizes the role of polygenic predisposition in PSE among stroke survivors. A PRS approach can be used to identify individuals at increased PSE risk and facilitate early intervention and preventive strategies. For instance, high-risk individuals could receive closer monitoring and targeted interventions, such as antiepileptic drugs, to prevent or mitigate the development of PSE.

Notably, the association between the PSE-specific PRS and PSE risk was specific to stroke/TIA survivors and not general epilepsy risk in the population. This finding further supports the notion of a unique polygenic predisposition to PSE in stroke/TIA survivors, emphasizing the significance of considering stroke history.

Our study has several strengths, including using large, diverse population-based samples from the UK Biobank and All of Us and identifying a PSE-specific PRS validated in an independent sample. However, our study also has a few limitations. Firstly, the ascertainment of PSE was based on diagnostic codes from electronic health records. It is possible that some cases of PSE were not captured, leading to an underestimation of the true incidence and prevalence of the condition. However, the use of electronic health records is a standard method for identifying PSE cases in large population-based studies, and the algorithms used to identify cases have been validated in previous studies.^29,30^ Secondly, we focused only on common genetic variants, and other rare genetic variants may also contribute to PSE development. Thirdly, stroke subtype data were unavailable for many UKB participants, and the instrument weights were obtained in participants with any stroke or TIA.

Future studies should investigate rare genetic variants associated with PSE and incorporate longitudinal data to investigate the temporal relationship between genetic risk factors and PSE development. Furthermore, studies investigating the biological mechanisms underlying the identified genetic variants’ association with PSE could provide insights into the PSE etiology and guide the development of targeted therapies. Additionally, the utility of PSE-specific PRS in predicting PSE risk should be evaluated in other populations and settings.

## CONCLUSION

Our study provides novel insights into the polygenic predisposition to PSE in stroke/TIA survivors and highlights the potential utility of PRS as a tool for predicting PSE risk. Our findings have important implications for developing personalized interventions and preventive strategies for PSE. Further research is needed to understand the mechanisms underlying PSE development better and identify additional genetic risk variants associated with this condition.

## ACKNOWLEDGMENTS

The authors would like to express their sincere gratitude to the All of Us Research Program participants and the UK Biobank participants, whose invaluable contributions made this study possible.

## DATA AVAILABILITY

The clinical data from the UK Biobank analyzed in our study is available to researcher on completion of the registration process (see https://www.ukbiobank.ac.uk/enable-your-research/register) and on successful application (see https://www.ukbiobank.ac.uk/enable-your-research/apply-for-access). The UK Biobank access committee approved this analysis as part of project 58743.

Data from the All of Us Research Program are publicly available at www.allofus.nih.gov. All study participants provided written informed consent. We used release version 7, which comprises data from all participants who enrolled from the beginning of the program on May 30, 2017, to June 23, 2022.

## POTENTIAL CONFLICTS OF INTEREST

All authors declare no financial relationships with any organizations that might have had an interest in the submitted work in the previous three years and no other relationships or activities that could appear to have influenced the submitted work.

## DISCLOSURES

Dr. de Havenon has received investigator-initiated clinical research funding from the NIH, consultant fees from Novo Nordisk, royalty fees from UpToDate, and equity in TitinKM and Certus. JZ reports speaker honoraria for unbranded educational events from UCB and Eisai, royalties/writer honoraria from Liber AB, Neurologi i Sverige, Studentlitteratur AB, and Wiley; as an employee of Sahlgrenska University Hospital being principal investigator/sub-investigator in clinical trials sponsored by Bial, SK Life science, GW Pharma and UCB (no personal compensation). NKM is an editorial board member of the *Neurology* and convenor of the IPSERC. Dr. Mazumder was supported by the Fogarty International Center of the National Institutes of Health under Award Number K01TW012178. The content is solely the responsibility of the authors and does not necessarily represent the official views of the National Institutes of Health.

---

**Figure 1.**
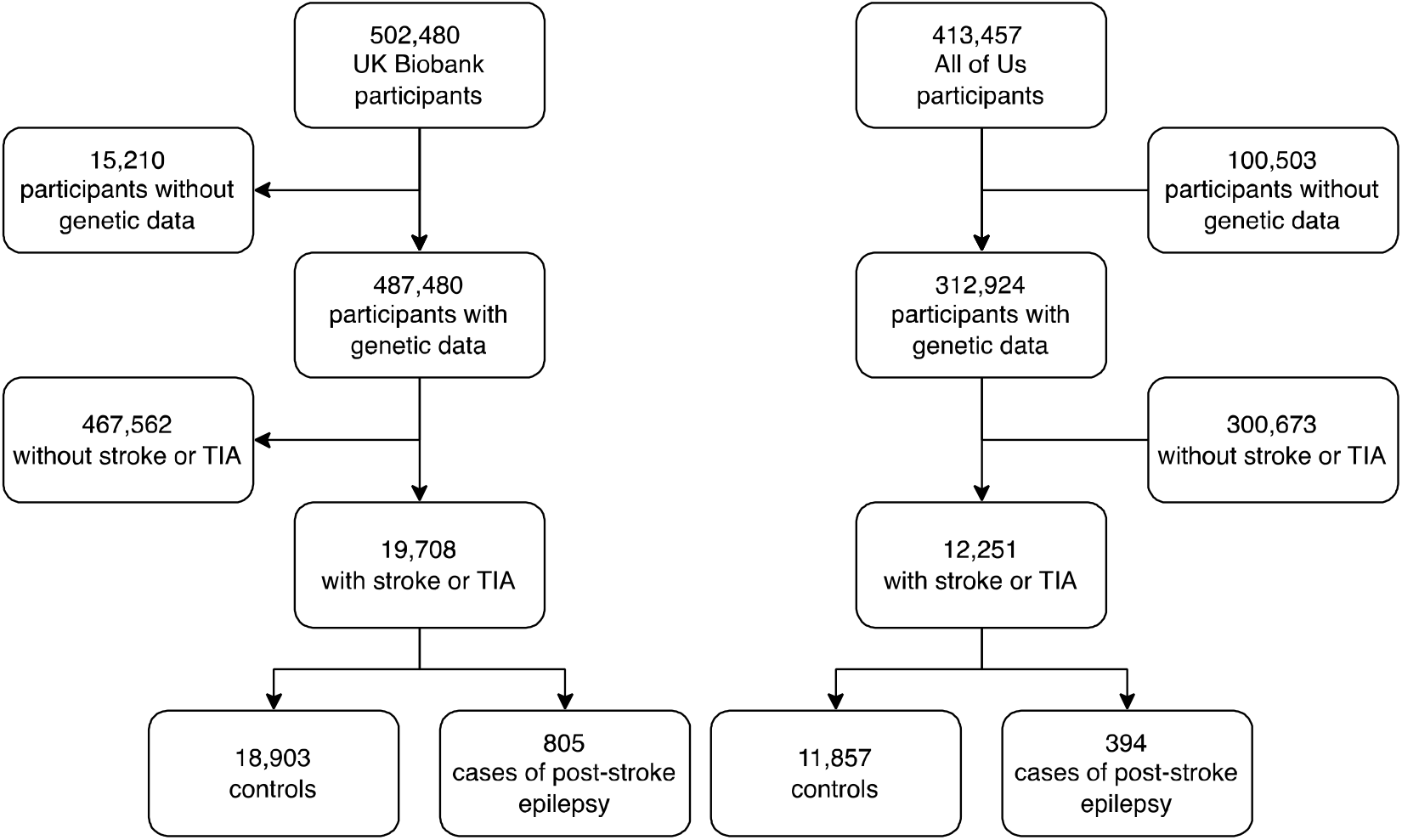
Flowchart of participants included in the analytical samples.

## Notes

### Competing Interest Statement

The authors have declared no competing interest.

### Author Declarations

The All of Us Research Program of the national Institutes of Health gave ethical approval for this work. The North West Centre for Research Ethics Committee gave ethical approval for this work.

