## Supplementary material for "Polygenic Risk of Epilepsy and Post-Stroke Epilepsy"

*** Jointly led this work.**

**** Jointly supervised this work.**

**Supplementary Table 1:** Standardized codes used to ascertain outcomes in the All of Us electronic health records (EHR).

| Condition | Source vocabulary | Source concept code | Source concept name |
|---|---|---|---|
| **Stroke (Ischemic)** | ICD10CM | G43.609 | Persistent migraine aura with cerebral infarction, not intractable, without status migrainosus |
| | ICD10CM | G43.619 | Persistent migraine aura with cerebral infarction, intractable, without status migrainosus |
| | ICD10CM | G46.5 | Pure motor lacunar syndrome |
| | ICD10CM | G46.6 | Pure sensory lacunar syndrome |
| | ICD10CM | G46.7 | Other lacunar syndromes |
| | ICD10CM | I63 | Cerebral infarction |
| | ICD10CM | P91.821 | Neonatal cerebral infarction, right side of brain |
| | ICD9CM | 347 | Persistent migraine aura with cerebral infarction, without mention of intractable migraine without mention of status migrainosus |
| | ICD9CM | 347 | Persistent migraine aura with cerebral infarction, with intractable migraine, so stated, without mention of status migrainosus |
| | ICD9CM | 433 | Occlusion and stenosis of basilar artery with cerebral infarction |
| | ICD9CM | 433 | Occlusion and stenosis of the carotid artery with cerebral infarction |
| | ICD9CM | 433 | Occlusion and stenosis of vertebral artery with cerebral infarction |
| | ICD9CM | 433 | Occlusion and stenosis of multiple and bilateral precerebral arteries with cerebral infarction |
| | ICD9CM | 434 | Occlusion and stenosis of other specified precerebral artery with cerebral infarction |
| | ICD9CM | 434 | Occlusion and stenosis of unspecified precerebral artery with cerebral infarction |
| | ICD9CM | 434 | Cerebral thrombosis with cerebral infarction |
| | ICD9CM | 434 | Cerebral embolism with cerebral infarction |
| | ICD9CM | 435 | Cerebral artery occlusion, unspecified with cerebral infarction |
| | SNOMED | 125081000119106 | Cerebral infarction due to occlusion of precerebral artery |
| | SNOMED | 149821000119103 | Cerebral infarction due to carotid artery occlusion |
| | SNOMED | 195185009 | Cerebral infarct due to thrombosis of precerebral arteries |
| | SNOMED | 195186005 | Cerebral infarction due to embolism of precerebral arteries |
| | SNOMED | 195189003 | Cerebral infarction due to thrombosis of cerebral arteries |
| | SNOMED | 195190007 | Cerebral infarction due to embolism of cerebral arteries |

| | | | |
|---|---|---|---|
| | SNOMED | 195230003 | Cerebral infarction due to cerebral venous thrombosis, non-pyogenic |
| | SNOMED | 230692004 | Infarction - precerebral |
| | SNOMED | 230698000 | Lacunar infarction |
| | SNOMED | 230702001 | Lacunar ataxic hemiparesis |
| | SNOMED | 230706003 | Hemorrhagic cerebral infarction |
| | SNOMED | 307767006 | Right sided cerebral infarction |
| | SNOMED | 34191000119104 | Cerebral infarction due to vertebral artery occlusion |
| | SNOMED | 426107000 | Acute lacunar infarction |
| | SNOMED | 432504007 | Cerebral infarction |
| | SNOMED | 433891000124100 | Cerebral infarction due to cerebral artery occlusion |
| | SNOMED | 705128004 | Cerebral infarction due to embolism of middle cerebral artery |
| | SNOMED | 705130002 | Cerebral infarction due to thrombosis of middle cerebral artery |
| | SNOMED | 99451000119105 | Cerebral infarction due to stenosis of carotid artery |
| **TIA** | ICD10CM | G45.0 | Vertebro-basilar artery syndrome |
| | ICD10CM | G45.8 | Other transient cerebral ischemic attacks and related syndromes |
| | ICD10CM | G45.9 | Transient cerebral ischemic attack, unspecified |
| | ICD9CM | 435 | Transient cerebral ischemia |
| | ICD9CM | 436 | Other specified transient cerebral ischemias |
| | ICD9CM | 436 | Unspecified transient cerebral ischemia |
| | SNOMED | 230717002 | Vertebrobasilar territory transient ischemic attack |
| | SNOMED | 266257000 | Transient cerebral ischemia |
| | SNOMED | 710575003 | Transient ischemic attack due to embolism |
| **Epilepsy** | ICD10CM | E88.42 | MERRF syndrome |
| | ICD10CM | G40.001 | Localization-related (focal) (partial) idiopathic epilepsy and epileptic syndromes with seizures of localized onset, not intractable, with status epilepticus |
| | ICD10CM | G40.009 | Localization-related (focal) (partial) idiopathic epilepsy and epileptic syndromes with seizures of localized onset, not intractable, without status epilepticus |

| | | | |
|---|---|---|---|
| | ICD10CM | G40.011 | Localization-related (focal) (partial) idiopathic epilepsy and epileptic syndromes with seizures of localized onset, intractable, with status epilepticus |
| | ICD10CM | G40.019 | Localization-related (focal) (partial) idiopathic epilepsy and epileptic syndromes with seizures of localized onset, intractable, without status epilepticus |
| | ICD10CM | G40.101 | Localization-related (focal) (partial) symptomatic epilepsy and epileptic syndromes with simple partial seizures, not intractable, with status epilepticus |
| | ICD10CM | G40.109 | Localization-related (focal) (partial) symptomatic epilepsy and epileptic syndromes with simple partial seizures, not intractable, without status epilepticus |
| | ICD10CM | G40.111 | Localization-related (focal) (partial) symptomatic epilepsy and epileptic syndromes with simple partial seizures, intractable, with status epilepticus |
| | ICD10CM | G40.119 | Localization-related (focal) (partial) symptomatic epilepsy and epileptic syndromes with simple partial seizures, intractable, without status epilepticus |
| | ICD10CM | G40.201 | Localization-related (focal) (partial) symptomatic epilepsy and epileptic syndromes with complex partial seizures, not intractable, with status epilepticus |
| | ICD10CM | G40.209 | Localization-related (focal) (partial) symptomatic epilepsy and epileptic syndromes with complex partial seizures, not intractable, without status epilepticus |
| | ICD10CM | G40.211 | Localization-related (focal) (partial) symptomatic epilepsy and epileptic syndromes with complex partial seizures, intractable, with status epilepticus |
| | ICD10CM | G40.219 | Localization-related (focal) (partial) symptomatic epilepsy and epileptic syndromes with complex partial seizures, intractable, without status epilepticus |
| | ICD10CM | G40.301 | Generalized idiopathic epilepsy and epileptic syndromes, not intractable, with status epilepticus |
| | ICD10CM | G40.309 | Generalized idiopathic epilepsy and epileptic syndromes, not intractable, without status epilepticus |
| | ICD10CM | G40.311 | Generalized idiopathic epilepsy and epileptic syndromes, intractable, with status epilepticus |
| | ICD10CM | G40.319 | Generalized idiopathic epilepsy and epileptic syndromes, intractable, without status epilepticus |
| | ICD10CM | G40.401 | Other generalized epilepsy and epileptic syndromes, not intractable, with status epilepticus |
| | ICD10CM | G40.409 | Other generalized epilepsy and epileptic syndromes, not intractable, without status epilepticus |
| | ICD10CM | G40.411 | Other generalized epilepsy and epileptic syndromes, intractable, with status epilepticus |
| | ICD10CM | G40.419 | Other generalized epilepsy and epileptic syndromes, intractable, without status epilepticus |
| | ICD10CM | G40.801 | Other epilepsy, not intractable, with status epilepticus |
| | ICD10CM | G40.803 | Other epilepsy, intractable, with status epilepticus |
| | ICD10CM | G40.811 | Lennox-Gastaut syndrome, not intractable, with status epilepticus |

| | | | |
|---|---|---|---|
| | ICD10CM | G40.813 | Lennox-Gastaut syndrome, intractable, with status epilepticus |
| | ICD10CM | G40.814 | Lennox-Gastaut syndrome, intractable, without status epilepticus |
| | ICD10CM | G40.821 | Epileptic spasms, not intractable, with status epilepticus |
| | ICD10CM | G40.823 | Epileptic spasms, intractable, with status epilepticus |
| | ICD10CM | G40.901 | Epilepsy, unspecified, not intractable, with status epilepticus |
| | ICD10CM | G40.91 | Epilepsy, unspecified, intractable |
| | ICD10CM | G40.911 | Epilepsy, unspecified, intractable, with status epilepticus |
| | ICD10CM | G40.919 | Epilepsy, unspecified, intractable, without status epilepticus |
| | ICD10CM | G40.A01 | Absence epileptic syndrome, not intractable, with status epilepticus |
| | ICD10CM | G40.A09 | Absence epileptic syndrome, not intractable, without status epilepticus |
| | ICD10CM | G40.A11 | Absence epileptic syndrome, intractable, with status epilepticus |
| | ICD10CM | G40.A19 | Absence epileptic syndrome, intractable, without status epilepticus |
| | ICD10CM | G40.B01 | Juvenile myoclonic epilepsy, not intractable, with status epilepticus |
| | ICD10CM | G40.B09 | Juvenile myoclonic epilepsy, not intractable, without status epilepticus |
| | ICD10CM | G40.B11 | Juvenile myoclonic epilepsy, intractable, with status epilepticus |
| | ICD10CM | G40.B19 | Juvenile myoclonic epilepsy, intractable, without status epilepticus |
| | ICD9CM | 345 | Generalized nonconvulsive epilepsy |
| | ICD9CM | 345 | Generalized nonconvulsive epilepsy, without mention of intractable epilepsy |
| | ICD9CM | 345 | Generalized nonconvulsive epilepsy, with intractable epilepsy |
| | ICD9CM | 345 | Generalized convulsive epilepsy |
| | ICD9CM | 345 | Generalized convulsive epilepsy, without mention of intractable epilepsy |
| | ICD9CM | 345 | Generalized convulsive epilepsy, with intractable epilepsy |
| | ICD9CM | 345 | Petit mal status |
| | ICD9CM | 345 | Grand mal status |
| | ICD9CM | 345 | Localization-related (focal) (partial) epilepsy and epileptic syndromes with complex partial seizures |

| | | | |
|---|---|---|---|
| | ICD9CM | 345 | Localization-related (focal) (partial) epilepsy and epileptic syndromes with complex partial seizures, without mention of intractable epilepsy |
| | ICD9CM | 345 | Localization-related (focal) (partial) epilepsy and epileptic syndromes with complex partial seizures, with intractable epilepsy |
| | ICD9CM | 346 | Localization-related (focal) (partial) epilepsy and epileptic syndromes with simple partial seizures |
| | ICD9CM | 346 | Localization-related (focal) (partial) epilepsy and epileptic syndromes with simple partial seizures, without mention of intractable epilepsy |
| | ICD9CM | 346 | Localization-related (focal) (partial) epilepsy and epileptic syndromes with simple partial seizures, with intractable epilepsy |
| | ICD9CM | 346 | Epilepsia partialis continua, with intractable epilepsy |
| | ICD9CM | 346 | Other forms of epilepsy and recurrent seizures, with intractable epilepsy |
| | ICD9CM | 346 | Epilepsy, unspecified, with intractable epilepsy |
| | SNOMED | 117891000119100 | Simple partial seizure |
| | SNOMED | 119001000119108 | Intractable simple partial epilepsy |
| | SNOMED | 13847000 | Partial seizure with impaired consciousness |
| | SNOMED | 13973009 | Grand mal status |
| | SNOMED | 192979009 | Generalized non-convulsive epilepsy |
| | SNOMED | 192992007 | Epileptic seizures - myoclonic |
| | SNOMED | 192999003 | Partial epilepsy with impairment of consciousness |
| | SNOMED | 193000002 | Temporal lobe epilepsy |
| | SNOMED | 193022009 | Localization-related(focal)(partial)idiopathic epilepsy and epileptic syndromes with seizures of localized onset |
| | SNOMED | 19598007 | Generalized epilepsy |
| | SNOMED | 230381009 | Localization-related epilepsy |
| | SNOMED | 230390002 | Localization-related symptomatic epilepsy |
| | SNOMED | 230394006 | Frontal lobe epilepsy |
| | SNOMED | 230426003 | Myoclonic epilepsy with ragged red fibers |
| | SNOMED | 230444006 | Menstrual epilepsy |
| | SNOMED | 230456007 | Status epilepticus |
| | SNOMED | 278510009 | Localization-related idiopathic epilepsy |

| | | | |
|---|---|---|---|
| | SNOMED | 290741000119102 | Intractable idiopathic partial epilepsy |
| | SNOMED | 29753000 | Partial seizure |
| | SNOMED | 3371000119106 | Refractory generalized convulsive epilepsy |
| | SNOMED | 352818000 | Tonic-clonic epilepsy |
| | SNOMED | 361123003 | Psychomotor epilepsy |
| | SNOMED | 36803009 | Idiopathic generalized epilepsy |
| | SNOMED | 37356005 | Myoclonic seizure |
| | SNOMED | 407675009 | Complex partial epileptic seizure |
| | SNOMED | 4103001 | Complex partial seizure with impairment of consciousness |
| | SNOMED | 422724001 | Refractory localization-related epilepsy |
| | SNOMED | 422873003 | Refractory epilepsia partialis continua |
| | SNOMED | 441678004 | Refractory generalized nonconvulsive epilepsy |
| | SNOMED | 442481002 | Epilepsy characterized by intractable complex partial seizures |
| | SNOMED | 445355009 | Refractory epilepsy |
| | SNOMED | 460731000124105 | Recurrent seizure |
| | SNOMED | 51075009 | Complex partial seizure evolving to generalized seizure |
| | SNOMED | 6204001 | Juvenile myoclonic epilepsy |
| | SNOMED | 65120008 | Generalized convulsive epilepsy |
| | SNOMED | 7033004 | Petit mal status |
| | SNOMED | 72103000 | Simple partial seizure with special sensory symptoms |
| | SNOMED | 74737003 | Complex partial seizure + impairment consciousness at onset |
| | SNOMED | 79348005 | Simple partial seizure, consciousness not impaired |
| | SNOMED | 79631006 | Absence seizure |
| | SNOMED | 79745005 | Reflex epilepsy |
| | SNOMED | 87095001 | Olfactory seizure |

**Supplemental Table 2:** Single Nucleotide Polymorphisms selected as genetic instruments including the PSE specific weights

| SNP | Trait | Chromosome | Base-pair position | Effect allele | MAF | Beta Cross | P Cross | Beta EUR | P EUR |
|---|---|---|---|---|---|---|---|---|---|
| rs1046276 | Juvenile myoclonic epilepsy | 16 | 30903305 | C | 0.34 | -0.07 | 0.2 | -0.08 | 0.12 |
| rs12185644 | Childhood absence epilepsy | 2 | 57824634 | A | 0.29 | -0.01 | 0.81 | 0 | 0.98 |
| rs13020210 | Childhood absence epilepsy | 2 | 144623658 | A | 0.2 | 0.04 | 0.56 | 0.03 | 0.71 |
| rs7587026 | Mesial temporal lobe epilepsy with hippocampal sclerosis | 2 | 166122240 | A | 0.263 | -0.05 | 0.36 | -0.05 | 0.4 |
| rs2947349 | Epilepsy | 2 | 57832668 | A | 0.26 | 0.06 | 0.29 | 0.07 | 0.21 |
| rs1939012 | Epilepsy | 11 | 102724404 | C | 0.4 | -0.01 | 0.82 | -0.01 | 0.84 |
| rs1044352 | Epilepsy | 4 | 31146252 | T | 0.5 | -0.02 | 0.7 | -0.02 | 0.66 |
| rs4665630 | Generalized epilepsy | 2 | 23675447 | T | 0.13 | 0.17 | 0.04 | 0.18 | 0.04 |
| rs1402398 | Generalized epilepsy | 2 | 57815106 | A | 0.36 | 0.05 | 0.31 | 0.06 | 0.27 |
| rs887696 | Generalized epilepsy | 2 | 190718781 | T | 0.34 | 0.02 | 0.73 | 0.02 | 0.78 |
| rs1044352 | Generalized epilepsy | 4 | 31146252 | T | 0.42 | -0.02 | 0.7 | -0.02 | 0.66 |
| rs11943905 | Generalized epilepsy | 4 | 46395600 | T | 0.27 | 0.04 | 0.43 | 0.04 | 0.44 |
| rs4596374 | Generalized epilepsy | 5 | 114885808 | T | 0.45 | -0.05 | 0.38 | -0.05 | 0.35 |
| rs68082256 | Generalized epilepsy | 6 | 16971344 | A | 0.2 | -0.05 | 0.41 | -0.06 | 0.35 |
| rs13200150 | Generalized epilepsy | 6 | 127988623 | G | 0.3 | 0 | 0.99 | 0.01 | 0.82 |
| rs4794333 | Generalized epilepsy | 17 | 47968129 | C | 0.38 | -0.05 | 0.32 | -0.06 | 0.23 |
| rs2833098 | Generalized epilepsy | 21 | 30811678 | A | 0.38 | 0.06 | 0.27 | 0.06 | 0.24 |
| rs36067110 | Generalized epilepsy | 1 | 17376448 | A | NR | -0.54 | 0.05 | -0.5 | 0.06 |
| rs34018214 | Generalized epilepsy | 1 | 17376363 | A | NR | -0.66 | 0.02 | -0.65 | 0.03 |
| rs1991545 | Focal epilepsy (with hippocampal sclerosis) | 3 | 155834522 | A | 0.04 | 0.13 | 0.4 | 0.08 | 0.64 |
| rs1318322 | Focal epilepsy (with hippocampal sclerosis) | 6 | 121013624 | G | 0.14 | 0.09 | 0.19 | 0.12 | 0.1 |
| rs2212656 | Focal epilepsy | 2 | 166144333 | A | 0.26 | -0.06 | 0.35 | -0.04 | 0.52 |

| | | | | | | | | | |
|---|---|---|---|---|---|---|---|---|---|
| rs12554609 | Focal epilepsy | 9 | 130447940 | G | NR | 0.09 | 0.29 | 0.08 | 0.33 |
| rs13026414 | Generalized epilepsy | 2 | 57706920 | T | 0.576 | 0 | 0.96 | -0.01 | 0.83 |
| rs11890028 | Generalized epilepsy | 2 | 166086767 | G | 0.688 | -0.03 | 0.6 | -0.03 | 0.63 |
| rs72823592 | Generalized epilepsy | 17 | 48045642 | A | 0.753 | -0.03 | 0.65 | -0.02 | 0.73 |
| rs10496964 | Generalized epilepsy | 2 | 144602342 | T | 0.844 | -0.03 | 0.7 | -0.02 | 0.75 |
| rs12059546 | Generalized epilepsy | 1 | 239806797 | G | 0.172 | -0.1 | 0.13 | -0.09 | 0.17 |
| rs6732655 | Epilepsy | 2 | 166038556 | T | 0.78 | 0.08 | 0.18 | 0.07 | 0.23 |
| rs28498976 | Epilepsy | 4 | 31149735 | A | 0.46 | -0.02 | 0.67 | -0.02 | 0.67 |
| rs4671319 | Epilepsy | 2 | 57723211 | A | 0.44 | 0 | 0.99 | 0 | 0.95 |
| rs6432877 | Epilepsy | 2 | 166142257 | G | 0.26 | -0.05 | 0.4 | -0.03 | 0.57 |
| rs4638568 | Epilepsy | 16 | 50011928 | A | 0.06 | -0.04 | 0.69 | -0.03 | 0.76 |
| rs2292096 | Epilepsy | 1 | 200857641 | G | NR | -0.02 | 0.75 | 0 | 0.98 |
| rs7139170 | Epilepsy | 12 | 112825713 | C | NR | -0.05 | 0.42 | -0.04 | 0.53 |
| rs11890028 | Epilepsy | 2 | 166086767 | G | NR | -0.03 | 0.6 | -0.03 | 0.63 |
| rs11978015 | Epilepsy | 7 | 86348656 | A | NR | -0.01 | 0.91 | 0 | 0.93 |
| rs28634186 | Epilepsy | 8 | 9811825 | C | NR | 0.03 | 0.64 | 0.01 | 0.87 |
| rs187977998 | Epilepsy | 4 | 62016346 | T | NR | 0.05 | 0.82 | 0.07 | 0.75 |

**Abbreviations-** SNP: Single Nucleotide Polymorphisms; MAF: Minor Allele Frequency; EUR: European ancestry; NR: Not Reported; Cross: Cross ancestry.

PSE-specific weights are given in Beta cross and Beta EUR columns.

**Supplemental Table 3:** Odds Ratios of developing PSE for one standard deviation increase in the PSE-specific PRS excluding participants with TIA.

| **Model** | **All of Us (3rd stage)** | | | |
|---|---|---|---|---|
| | All ancestries | | European ancestry | |
| | OR (95% CI) | P-value | OR (95% CI) | P-value |
| **Univariable** | 1.13 (1.01-1.26) | 0.028 | 1.14 (0.91-1.43) | 0.244 |
| **Multivariable*** | 1.18 (1.02-1.38) | 0.031 | 1.15 (0.91-1.44) | 0.228 |

**Abbreviations-** OR: Odds Ratio; CI: Confidence Interval.
*Adjusted for age, sex, genetic ancestry, and the first four genetic principal components.

**Supplemental Table 4:** Characteristics of the two cohorts restricted to European ancestry.

| | UK Biobank | All of Us |
|---|---|---|
| **Baseline demographics** | | |
| Number of participants (n) | 16,903 | 5,953 |
| Age (mean (SD)) | 61.33 (6.47) | 67.75 (12.10) |
| Female sex | 9,699 (57.4%) | 3,004 (50.5%) |
| **Events of interest** | | |
| Stroke | 10,803 (63.9%) | 3,714 (62.4%) |
| TIA | 8,167 (48.3) | 3,359 (56.4%) |
| Epilepsy | 1,161 (6.9%) | 305 (5.1%) |
| Post-stroke epilepsy | 705 (4.2%) | 155 (2.6%) |

**Abbreviations-** SD: Standard Deviation; TIA: Transient Ischemic Attack.
